# Identification of a novel FUS/ETV4 fusion and comparative analysis with other Ewing sarcoma fusion proteins

**DOI:** 10.1101/2021.05.07.21256326

**Authors:** Megann A. Boone, Cenny Taslim, Jesse C. Crow, Julia Selich-Anderson, Mike Watson, Peter Heppner, James Hamill, Andrew C. Wood, Stephen L. Lessnick, Mark Winstanley

## Abstract

Ewing sarcoma is an aggressive pediatric bone cancer defined by a chromosomal translocation fusing one of the FET family members to a member of the ETS transcription factor family. To date, there have been seven reported translocations, with the most recent translocation reported over a decade ago. We now report the first identification of a novel translocation occurring between the *FUS* gene and ETS family member *ETV4* detected in a neonatal patient with Ewing sarcoma. Given its apparent rarity, we conducted an initial characterization of FUS/ETV4 function by performing genomic localization and transcriptional regulatory studies. We knocked down endogenous EWS/FLI in the A673 cell line, and expressed FUS/ETV4 in its stead, and performed CUT&Tag and RNA-sequencing analyses. We compared these data to similar “knock-down/rescue” analyses of other rare (non-EWS/FLI) Ewing sarcoma-associated translocation products. Through this comparative analysis in the same genetic background, we demonstrate significant similarities across these fusions, and in doing so, validate this novel FUS/ETV4 translocation as a *bona fide* Ewing sarcoma translocation. This study presents the first genomic comparisons of the rare Ewing sarcoma-associated translocation products, and reveals that the FET/ETS fusions share highly similar, but not identical, genomic localization and transcriptional regulation patterns. These data provide insights into the roles of both the FET and ETS sides of these fusions, and provide a generic strategy to provide further strength to the notion that FET/ETS fusions are key drivers of, and thus pathognomonic for, Ewing sarcoma.

**Significance:** Identification and initial characterization of the novel Ewing sarcoma fusion, FUS/ETV4, expands the family of Ewing-fusions and extends the diagnostic possibilities for this aggressive tumor of adolescents and young adults.

## Introduction

Ewing sarcoma is an aggressive bone- and soft tissue-associated cancer primarily diagnosed in children and young adults (1, 2). The disease is characterized by the presence of a chromosomal translocation that encodes fusions between the amino-terminal domain of a FET (<u>F</u>US, <u>E</u>WSR1, and <u>T</u>AF15) protein to the carboxyl-terminal domain of an ETS (<u>E</u>26 <u>T</u>ransformation-<u>S</u>pecific) transcription factor family member. The most common chromosomal translocation, present in ∼85% of cases, is the t(11;22)(q24:q12), that fuses the *EWSR1* gene to *FLI1* to encode the EWS/FLI fusion oncoprotein (1, 2). EWS/FLI functions as an aberrant transcription factor that uses its ETS domain to bind DNA and the EWS-portion to regulate gene expression. Following the identification of EWS/FLI nearly three decades ago, an EWS/ERG fusion was found in ∼10% of cases, followed by five other fusions that are present in <1% of cases each: EWS/FEV, EWS/ETV1, EWS/ETV4, FUS/ERG, and FUS/FEV (3, 4). Each of these is believed to function as an aberrant transcription factor, primarily on the basis of their similar domain structure to EWS/FLI. Here, we report a novel eighth Ewing sarcoma fusion, FUS/ETV4, identified in a neonatal patient.

The ETS protein family is a large group of transcription factors characterized by a highly-conserved DNA-binding domain, with structural variability outside of this region contributing to subfamily classification (5). The five ETS members identified in Ewing sarcoma fusions derive from two of these subfamilies: FLI, ERG, and FEV are members of the ERG subfamily, and ETV1 and ETV4 of the PEA3 subfamily (1, 2, 5). It is believed that these ETS family members bind similar high-affinity target sequences in vitro, but whether they have similar genomic localization in the context of the Ewing sarcoma fusions is unknown. This is of particular interest given the neomorphic capability of EWS/FLI to bind and regulate genes via GGAA-microsatellites in the human genome.

The amino-terminal intrinsically-disordered regions (IDRs) of EWS and FUS have biophysical features that appear critical to the ability of FET/ETS proteins to bind DNA and regulate gene expression. These IDRs have self-association properties that mediate phase separation and/or “hub” formation (6, 7). These unique properties are likely critical for FET/ETS fusion oncoprotein-mediated reorganization of chromatin architecture, formation of transcriptional hubs, and recruitment of necessary transcriptional co-factors, such as the BAF complex, and are thus likely crucial for Ewing sarcomagenesis (1, 2).

The vast majority of molecular studies of Ewing sarcoma fusions have focused on EWS/FLI, but there are few, if any, detailed analyses of other fusion proteins. While it makes logical sense that FET/ETS fusions will have similar biologic functions, this has not been formally demonstrated. This focus on EWS/FLI and relative lack-of-focus on the other Ewing fusions has significant impact on the clinical management of patients (8). The advent of next-generation sequencing has allowed for the ready identification of EWS-based and FUS-based fusion transcripts or genomic-rearrangements (9). These technological advances have made the identification of fusion partners easier, and simultaneously raised new questions as to how to apply this information to clinical care. As an example, a recent survey by the Children’s Oncology Group found that only ∼35% of clinician respondents indicated that non-EWS/FLI FUS/ETS fusions should be classified as Ewing sarcoma (8). Importantly, a significant portion of respondents indicated they were unsure whether alternative FET/ETS fusions should even be used to diagnose Ewing sarcoma or to allow patients to be included in Ewing sarcoma clinical trials (8).

We now report the initial identification of a novel FUS/ETV4 fusion in a patient with Ewing sarcoma, and perform genomic localization and transcriptional studies in an Ewing sarcoma A673 knock-down/rescue model system. We used this same system to perform the first comparative analysis of other rare, non-EWS/FLI, fusions in Ewing sarcoma, and in doing so, we demonstrate strong similarities between all of the fusions, demonstrating that they are indeed functionally similar. At the same time, we find some differences between FET/ETS fusions that might represent differences in DNA binding function and interplay between the FET-and ETS-portions of the fusions. These data support the conclusion that all FET/ETS translocations should be regarded as *bona fide* Ewing sarcoma translocations and clinically classified as such.

## Materials and Methods

### Constructs and retroviruses

Puromycin-resistant retroviral vectors encoding shRNAs targeting Luciferase (iLuc; sequence: 5’-GATCCCCCTTACGCTGAGTACTTCGATTCAAGAGATCGAAGTACTCAGCGTAAGTTTTTGG AAC-3”) or the 3’-UTR of endogenous EWS/FLI mRNA (iEF; sequence: 5’-GATCCCCATAGAGGTGGGAAGCTTATTTCAAGAGAATAAGCTTCCCACCTCTATTTTTTGGAAC-3’) were previously described (10, 11). FET/ETS fusions (containing amino-terminal 3xFLAG-tags) were cloned into pMSCV-Hygro (Invitrogen); sequence details provided in Supplementary Table 1.

### Cell culture methods

HEK-293EBNA (Invitrogen) and A673 cells (ATCC), cultured for 1-6 weeks, in appropriate media and retroviruses produced and used for infection as described (10-12). STR profiling and mycoplasma testing are performed annually on all cell lines.

### Immunodetection

Whole-cell protein extraction, protein quantification, and Western blot analysis was performed as previously described (10-12). Immunoblotting was performed using anti-FLAG M2 mouse (Sigma F1804-200UG) and anti-α-Tubulin (Abcam ab7291). Membranes were imaged using the LiCor Odyssey CLx Infrared Imaging System.

### qRT-PCR

Total RNA was extracted from cells using the RNeasy Extraction Kit (Qiagen 74136). Reverse transcription and qPCR were performed using the iTaq Universal SYBR Green 1-Step Reaction Mix (BioRad 1725151) on a Bio-Rad CFX Connect Real-Time System. Primer sequences listed in Supplementary Table 2.

### CUT&Tag and Analysis

CUT&Tag (Cleavage Under Targets and Tagmentation) was performed as described by (13) on two biological replicates of knock-down/rescue A673 samples using the anti-FLAG M2 mouse antibody (1:100, Sigma F1804-200UG), and sequenced on the Illumina HiSeq4000 platform. Reads were trimmed, de-duplicated using SAMTOOLS (RRID:SCR_002105), aligned to hg19 reference genome, spike-in normalized using DESeq2 (median ratio method, RRID:SCR_015687), tracks were generated and averaged across biological replicates using Deeptools (RRID:SCR_016366), and peaks were called using MACS (RRID:SCR_013291), DiffBind (RRID:SCR_012918), and DESeq2 (14-16). Peaks were called as significant with the following parameters: Irreproducible Discovery Rate of 0.01, FDR (False Discovery Rate) < 0.05, log2(fold-change) > 3 over control samples (iEF+Empty Vector), mean normalized counts > 80. Overlaps were determined using VennDiagram (RRID:SCR_002414) and GenomicRanges (RRID:SCR_000025) (17).

### RNA-sequencing and Analysis

RNA-sequencing was performed on two biological replicates of knock-down/rescue A673 cell samples. TruSeq Stranded mRNA Kit (Illumina Cat. No. 20020594) was used to prepare cDNA libraries from total RNA and sequenced on Illumina HiSeq4000 to generate 150-bp paired-end reads. Reads were analyzed for quality control, trimmed, aligned to the human genome and analyzed for differential expression using FastQC (RRID:SCR_014583), MultiQC (RRID:SCR_014982), Trim_Galore (RRID:SCR_011847), STAR (RRID:SCR_004463, version b), and DESeq2 (16). Venn diagrams were created for differentially expressed genes for samples compared to control cells (iEF+Empty Vector) (FDR < 0.05).

### Statistical Analysis

PCR data is presented as mean ± SEM. Significance of soft agar assays was determined using a Student’s t-test, or as otherwise noted; p-values < 0.05 were considered to be significant.

## Results & Discussion

### Identification of a novel FUS/ETV4 translocation

An infantile patient presented with a left posterior mediastinal mass (Figure 1A). The mass occupied a significant portion of the left thoracic cavity and extensive intraspinal extension was observed from T3 to T8 without evidence of metastatic disease. A thoracic laminoplasty and resection of the intraspinal component was performed to manage the severely compressed spinal cord. The pathology of open biopsy specimens revealed classic Ewing sarcoma with sheets of small round blue-staining cells with no evidence of differentiation (Figure 1B). The tumor was CD99-positive in a diffuse membranous staining pattern (Figure 1C), and positive for nuclear NKX2-2 expression (Figure 1D). *EWS* rearrangement was not detected, so *FUS* break-apart FISH was performed and identified a rearrangement. Commercial molecular genetic testing revealed a translocation between the *FUS* locus on chromosome 16p11.2 and the *ETV4* locus on chromosome 17q21. This translocation encoded an in-frame fusion between exons 1-9 of *FUS* to exons 10-13 of *ETV4*. A literature search revealed the FUS/ETV4 translocation to be a novel fusion, previously unreported and undiscussed.

**Figure 1.**
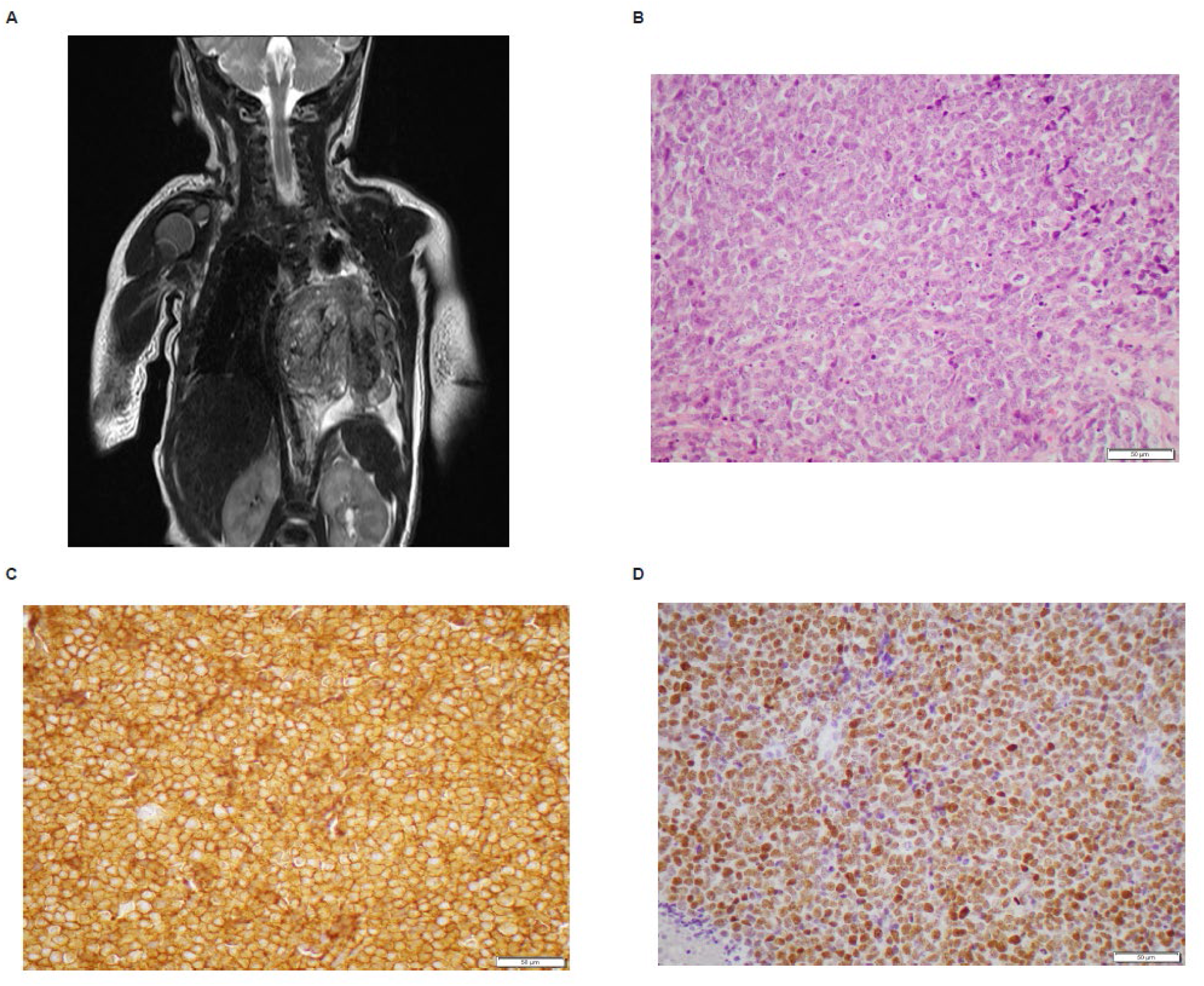
Neonatal patient presenting with Ewing sarcoma tumor. (A) Coronal magnetic resonance imaging (MRI) scan revealed a left posterior mediastinal mass. (B) Hematoxylin and eosin staining of patient tumor biopsy revealed sheets of undifferentiated, mitotically active small, round blue cells with dispersed chromatin and minimal amphophilic cytoplasm (50 μM scale bar depicted on image). (C) CD99 immunochemistry reveals diffuse membranous expression (50 μM scale bar depicted on image). (D) NKX2-2 immunohistochemistry shows diffuse strong nuclear immunoreactivity (50 μM scale bar depicted on image).

### FUS/ETV4 has similar binding and transcriptional functions to EWS/ETV4

There were no cell lines nor patient-derived xenograft models available from the patient in which to analyze the transcriptional functions of FUS/ETV4. We therefore cloned FUS/ETV4 into a retroviral expression vector, and also cloned EWS/ETV4 as the most similar *bona fide* rare Ewing sarcoma fusion (Figure 2A). To allow for analysis in an isogenic background, we knocked-down endogenous EWS/FLI in A673 Ewing sarcoma cells and expressed either EWS/ETV4 or FUS/ETV4 fusion proteins through retroviral transduction (Supplemental Figure 1A-B).

**Figure 2.**
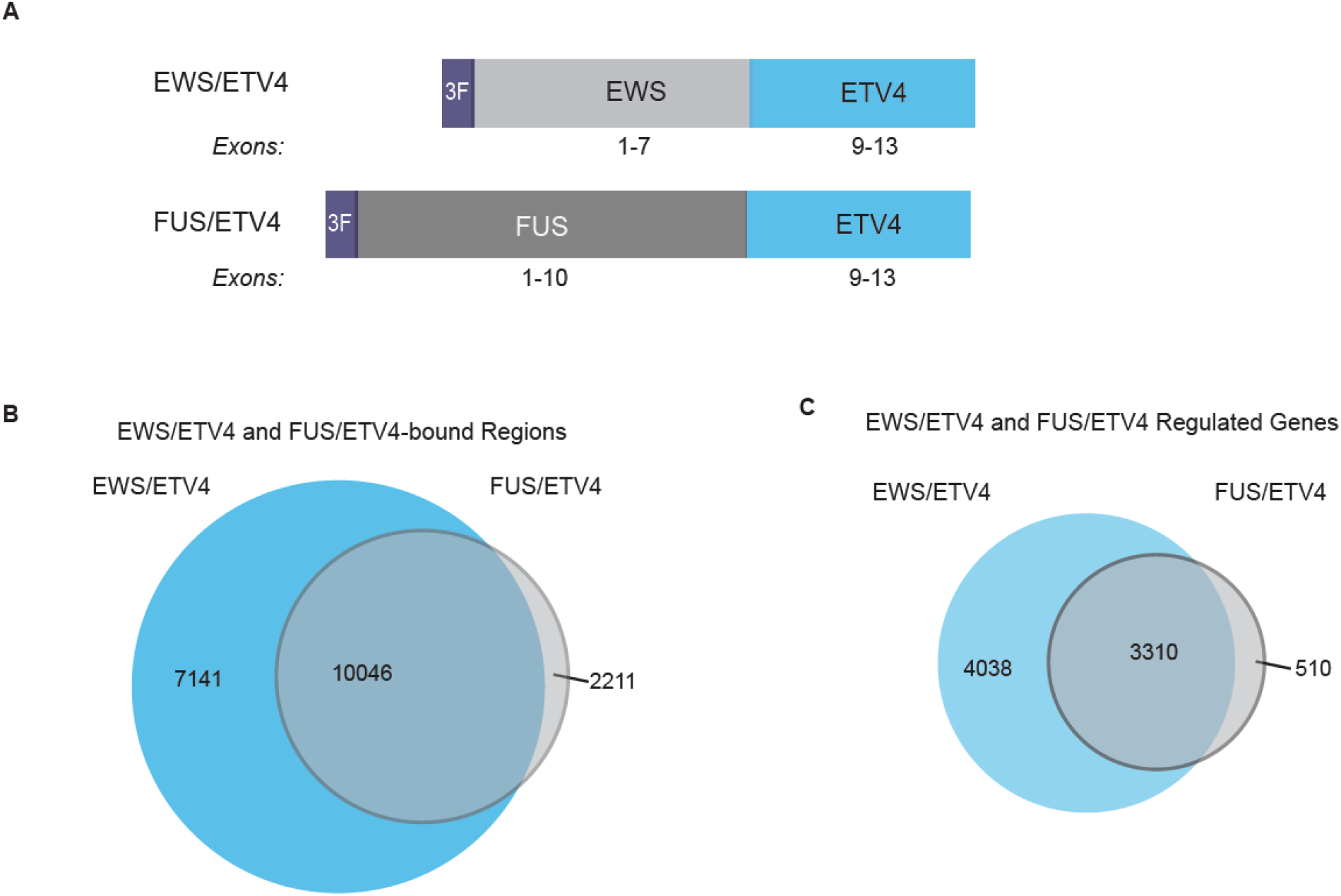
EWS/ETV4 and FUS/ETV4 DNA-binding and transcriptional profile overlap reveals similar biological functions. (A) Protein schematic of 3xFLAG-tagged (3F) EWS/ETV4 and FUS/ETV4 constructs. EWS is represented in light grey, FUS in dark grey, and ETV4 in light blue. Exons included in each fusion are noted. (B) Venn diagram overlap analysis performed on CUT&Tag-detected genomic localization data for EWS/ETV4 and FUS/ETV4 expressed in A673 knock-down/rescue cells, as compared to control cells (Control: iEF + Empty Vector; EWS/ETV4: iEF + EWS/ETV4; FUS/ETV4: iEF + FUS/ETV4) (N=2). The number of peaks uniquely bound by each construct or those that are similarly bound are indicated in the figure. Significance of overlap: p < 2.2 x 10^−16^. (C) Venn diagram analysis of RNA-sequencing results depicting significantly regulated genes for EWS/ETV4 and FUS/ETV4-expressing A673 knock-down rescue cells, as compared to iEF + Empty Vector control cells (N=2). Number of regulated genes for each construct is indicated in the figure. Significance of overlap: p < 2.2 x 10^−16^.

We first compared genome-wide localization of FUS/ETV4 and EWS/ETV4 using CUT&Tag (13). Both constructs were 3xFLAG-tagged and the use of the same anti-FLAG antibody for genomic localization allowed the data to be compared directly, without the confounding effects of using different antibodies with different affinities and specificities. We found that FUS/ETV4 bound >12,000 loci and EWS/ETV4 bound >17,000 loci. Strikingly, >10,000 bound loci were shared between the two proteins, and over 80% of FUS/ETV4 peaks overlapped with those of EWS/ETV4 (Figure 2B).

We next asked whether FUS/ETV4 induced a similar transcriptional profile to EWS/ETV4. Consistent with the genomic localization studies, RNA-sequencing revealed that 87% of the genes regulated by FUS/ETV4 were also regulated by EWS/ETV4, although EWS/ETV4 again regulated more genes than FUS/ETV4 (Figure 2C). Both fusions were capable of binding and regulating genes previously documented as EWS/FLI targets, including those associated with both high-affinity and GGAA-microsatellite binding sites (Supplemental Figure 1C-D). Taken together, these data demonstrate that the novel FUS/ETV4 fusion has transcriptional function that are similar to EWS/ETV4, and thus supports its identity as a *bona fide* Ewing sarcoma fusion.

### ERG- and FEV-based fusions have similar binding and transcriptional functions

We recognized that the A673 knock-down/rescue system could be generalized to compare other understudied Ewing sarcoma fusion proteins, particularly those ETS-family members that have both EWS- and FUS-versions. We therefore compared EWS/ERG to FUS/ERG, and EWS/FEV to FUS/FEV (Figure 3A and Supplemental Figure 2A-B; NB: neither EWS/FLI nor EWS/ETV1 have FUS-versions identified to date). We found that almost 13,000 bound loci were shared between EWS/ERG and FUS/ERG, with >80% of the EWS/ERG loci also bound by FUS/ERG (Figure 3B, left panel; NB: The higher number of FUS/ERG-bound loci likely reflects higher protein expression of FUS/ERG, see Supplemental Figure 2B). Similarly, EWS/FEV and FUS/FEV shared almost 15,000 bound regions, accounting for ∼70% of the regions bound by both fusion proteins (Figure 3B, right panel).

**Figure 3.**
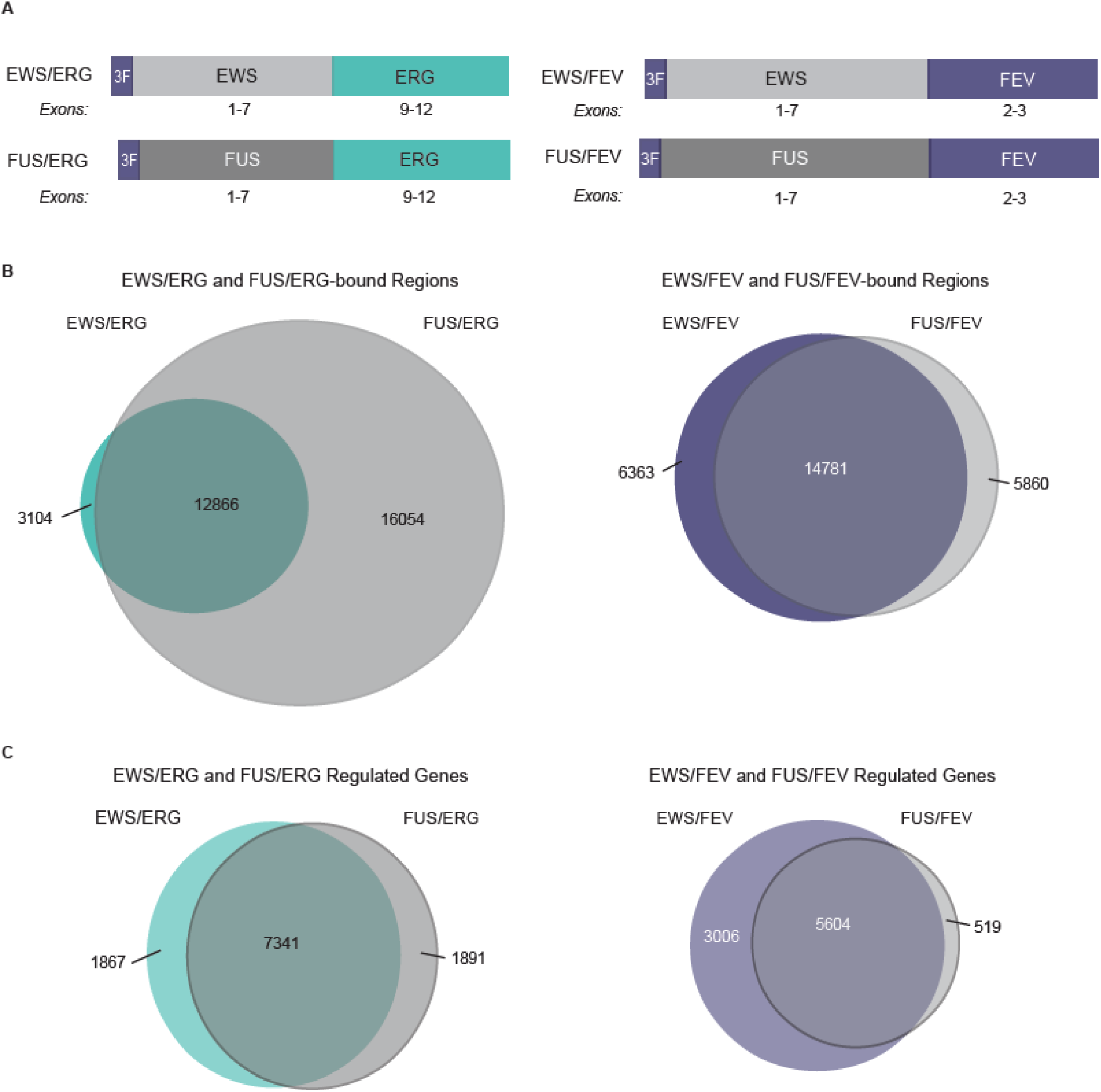
Comparison of FET/ERG and FET/FEV fusions shows similar genomic localization and transcriptional regulatory profiles. (A) Protein schematic of 3xFLAG-tagged (3F) cDNA constructs, including EWS/ERG, FUS/ERG, EWS/FEV, and FUS/FEV. EWS is depicted in light grey, FUS in dark grey, ERG in teal, and FEV in indigo. Exons included in each fusion are noted. (B) Venn diagram overlap analysis of CUT&Tag genomic localization data for the corresponding fusion protein listed after expression in A673 knock-down/rescue cells (iEF + Construct), as compared to control cells (iEF + Empty Vector) (N=2). Number of bound regions for each construct depicted in figure. Significance of overlap: p < 2.2 x 10^−16^. (C) Venn diagram overlap analysis of RNA-sequencing expression data for genes called as significantly regulated by the corresponding construct listed in A673 knock-down/rescue cells, as compared to control cells (iEF + Empty Vector) (N=2). Number of significantly regulated genes by each fusion listed in figure. Significance of overlap: p < 2.2 x 10^−16^.

RNA-sequencing revealed an ∼80% overlap between genes regulated by EWS/ERG and FUS/ERG and that each regulated >9,000 genes (suggesting that much of the “excess” FUS/ERG binding was not-functionally associated with gene regulation; Figure 3C, left panel). Similarly, EWS/FEV and FUS/FEV regulated ∼5,600 genes in common, representing ∼65% of the genes regulated by EWS/FEV and ∼90% of genes regulated by FUS/FEV (Figure 3C, right panel).

Taken together with the ETV4-fusion data above, the finding that EWS/ETS and FUS/ETS fusions bind similar loci and regulate similar sets of genes suggest that the EWS- and FUS-regions of the fusions are largely interchangeable, and strengthen the notion that tumors harboring these fusions should all be considered Ewing sarcomas.

### EWS- and FUS-based fusions have similar binding and transcriptional functions

The analyses above demonstrated that fusions with the same ETS domain bind and regulate gene expression in a similar manner regardless of whether the fusion partner is EWS or FUS. We next sought to determine if DNA binding and gene regulation would be similar in fusions that had the same amino-terminus (either EWS or FUS), but differed in their ETS domain. We compared EWS/ETV4, EWS/ERG, and EWS/FEV alongside EWS/FLI (the most common Ewing sarcoma fusion) as a group, and FUS/ETV4, FUS/ERG, and FUS/FEV as a group in the A673 knock-down/rescue system (Supplemental Figure 2A-B). The EWS/ETS fusions shared almost 9,000 bound loci (Supplemental Figure 3A), and the FUS/ETS fusions shared >8,700 bound loci (Supplemental Figure 3B). RNA-sequencing showed similar trends, with >5,400 genes similarly regulated by each of the EWS/ETS fusions (Supplemental Figure 3C), and ∼2,900 genes regulated by the FUS/ETS proteins (Supplemental Figure 3D).

### Global similarities across all FET/ETS fusions support the inclusion of all tumors harboring FET/ETS fusions as *bona fide* Ewing sarcomas

Lastly, we asked whether the similarities in DNA-binding and transcriptional regulation we observed in each “class” of fusion (grouped based on ETS domain or on amino-terminal domain) would be observed across the entire group of FET/ETS fusions included herein. The data generated above was therefore analyzed *in toto*. Genomic localization revealed that over 6,600 loci were similarly bound and that ∼2,600 genes were similarly regulated by all fusion proteins tested (Figure 4A-B). These overlaps were highly significant (p<2.2×10^−16^). We again observed that all fusions bound and regulated both GGAA-microsatellite associated genes, and genes associated with high-affinity ETS binding sites (Supplemental Figures 4A-B and 5A-B). Taken together, these data support the assertion that all FET/ETS fusion proteins have similar capabilities to bind DNA and regulate gene expression.

**Figure 4.**
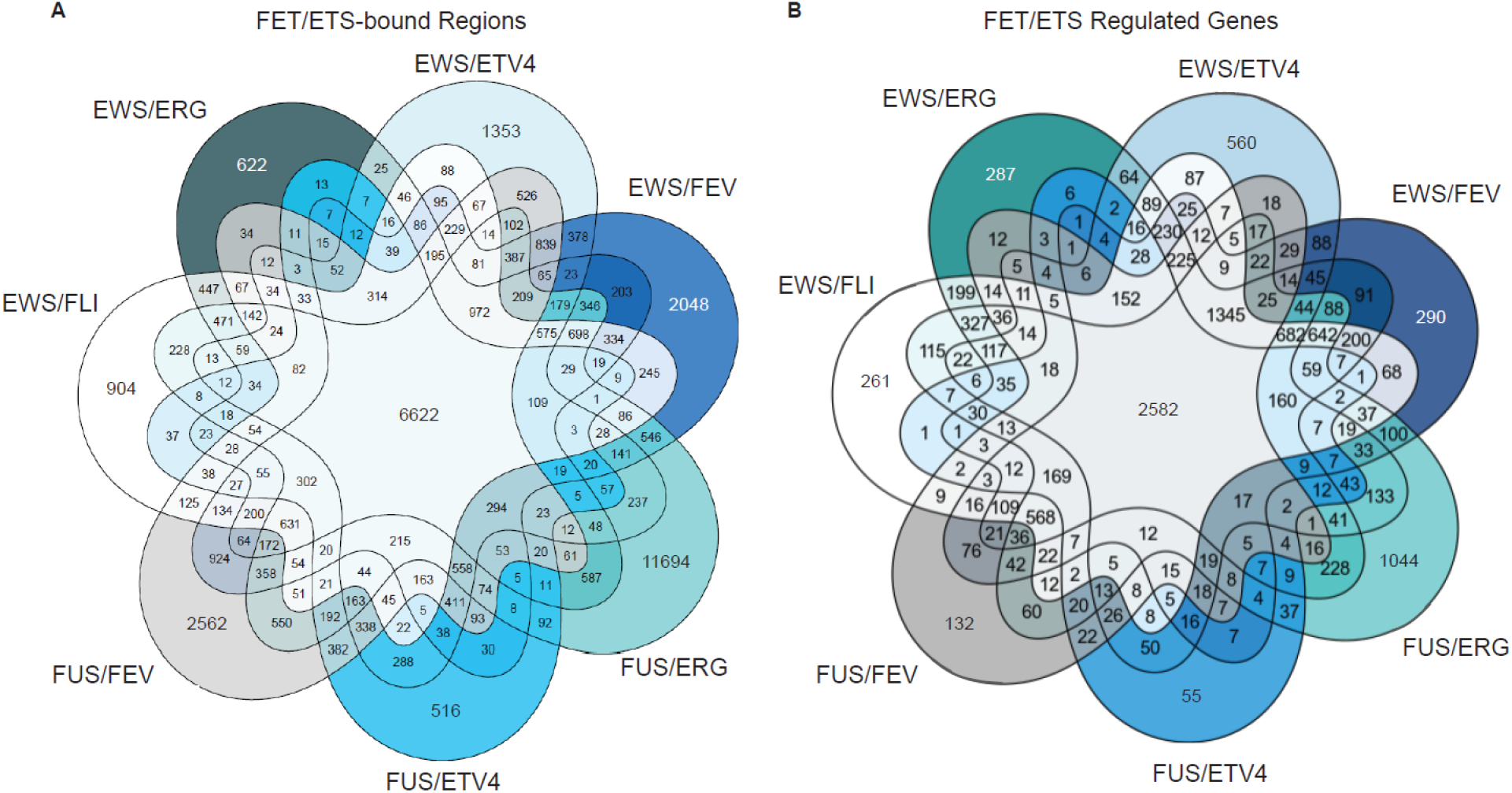
Analysis of DNA-bound regions and regulated genes by FET/ETS fusion proteins reveals significant overlap. (A) Venn diagram overlap analysis of CUT&Tag genomic localization binding data of FET/ETS translocations in A673 knock-down/rescue cells (N=2). All bound regions are called as significant for the corresponding translocation as compared to control cells (iEF + Empty Vector). Significance of overlap: p < 2.2 x 10^−16^. (B) Venn diagram analysis of significantly regulated genes by corresponding FET/ETS translocations, as compared to control cells (iEF + Empty Vector) determined using RNA-sequencing (N=2). Significance of overlap: p < 2.2 x 10^−16^.

The most common fusion in Ewing sarcoma, EWS/FLI, has been extensively studied (1, 2, 18). This work has led to development of novel concepts for EWS/FLI protein function, including the function of the EWS-portion of the fusion as a transcriptional regulatory domain, likely through the assembly of transcriptional hubs via self-association properties, the ability of the EWS-portion to recruit transcriptional co-regulators, such as BAF and LSD1, and the ability of the fusion to alter chromatin architecture (1, 2, 6, 19). Undergirding these properties lies the ability of the fusion to localize to specific loci in the genome, including those harboring GGAA-microsatellites and/or high-affinity ETS binding sites, and to dysregulate gene expression ultimately resulting in the formation of Ewing sarcoma. Although additional Ewing sarcoma translocations have been identified, the analysis of these fusions has been rudimentary at best and investigators have simply assumed similar function based on similar structure. At face value this seems reasonable, but leaves many unanswered questions, such as if functional differences in the fusions exist that might result in some being more rarely associated with Ewing sarcoma, or whether there is a critical interplay between fusion type and cellular background that is required for Ewing sarcoma development. Finally, the lack of important comparative analyses has allowed for confusion to arise in the clinical management of patients with likely Ewing sarcoma that harbor one of the rare translocations. Indeed, patients with rare translocations may not be offered entry onto clinical trials designed for patients with Ewing sarcoma and may therefore lead to subpar care (8).

In this report, we describe the identification of a novel FUS/ETV4 patient translocation. We demonstrate that this fusion shares many of the DNA-binding and gene regulatory properties of other Ewing sarcoma-associated fusion proteins, including the well-studied EWS/FLI fusion. Through a large-scale comparison between variant Ewing fusions in an isogenic system, we find that all of the Ewing fusions analyzed share significant similarities in DNA-binding and gene regulation. These data support the notion that the novel FUS/ETV4 fusion reported here is a *bona fide* Ewing sarcoma translocation, and suggest that FET/ETS translocations bind and regulate similar target genes to mediate oncogenesis. Accordingly, these data support that tumors containing FET/ETS translocations should be clinically diagnosed as Ewing sarcoma tumors and justifies the inclusion of patients with these tumors in standard and experimental Ewing sarcoma treatment protocols, as well as clinical trials.

## Data Availability

The sequencing datasets generated and analyzed during the current study are available in the Gene Expression Omnibus and accessible at GSE173185. All other data generated or analyzed during this study are available from the corresponding author upon reasonable request.

## Declarations

### Ethics

Nationwide Children’s Hospital Institutional Review Board determined that this project was not classified as human subjects research and was therefore exempt from review.

## Acknowledgements

We thank Dr. Susan Arbuckle, Children’s Hospital at Westmead, Sydney, Australia, for assistance with NKX2-2 staining of tumor samples. We also thank Dr. Andrea K. Byrum, Dr. Emily R. Theisen, Dr. Jack Tokarsky, Ariunaa Bayanjargal, and Iftekhar Showpnil for thoughtful discussion concerning the hypothesis and methodology of the project, as well as copyediting this manuscript. Additionally, we would like to acknowledge the generosity of the patient’s family for the use of the tumor sequencing information in this study.

## Authors Contributions

MAB, SLL, and MW(instanley) are responsible for conceptualization of the project. For patient care, MW(instanley) was acting oncologist, MW(atson) was acting pathologist, PH was acting neurosurgeon, JH was surgeon responsible for local tumor control, and AW provided patient sequencing guidance. Methodology for laboratory studies was formulated by MAB and SLL. Investigation was performed by MAB, JCC, and JSA. Data analysis was performed by MAB and CT. Manuscript preparation was completed by MAB and reviewing and editing was performed by all authors. Funding acquisition and supervision completed by SLL.

## Figure Legends

**Supplemental Figure 1.**
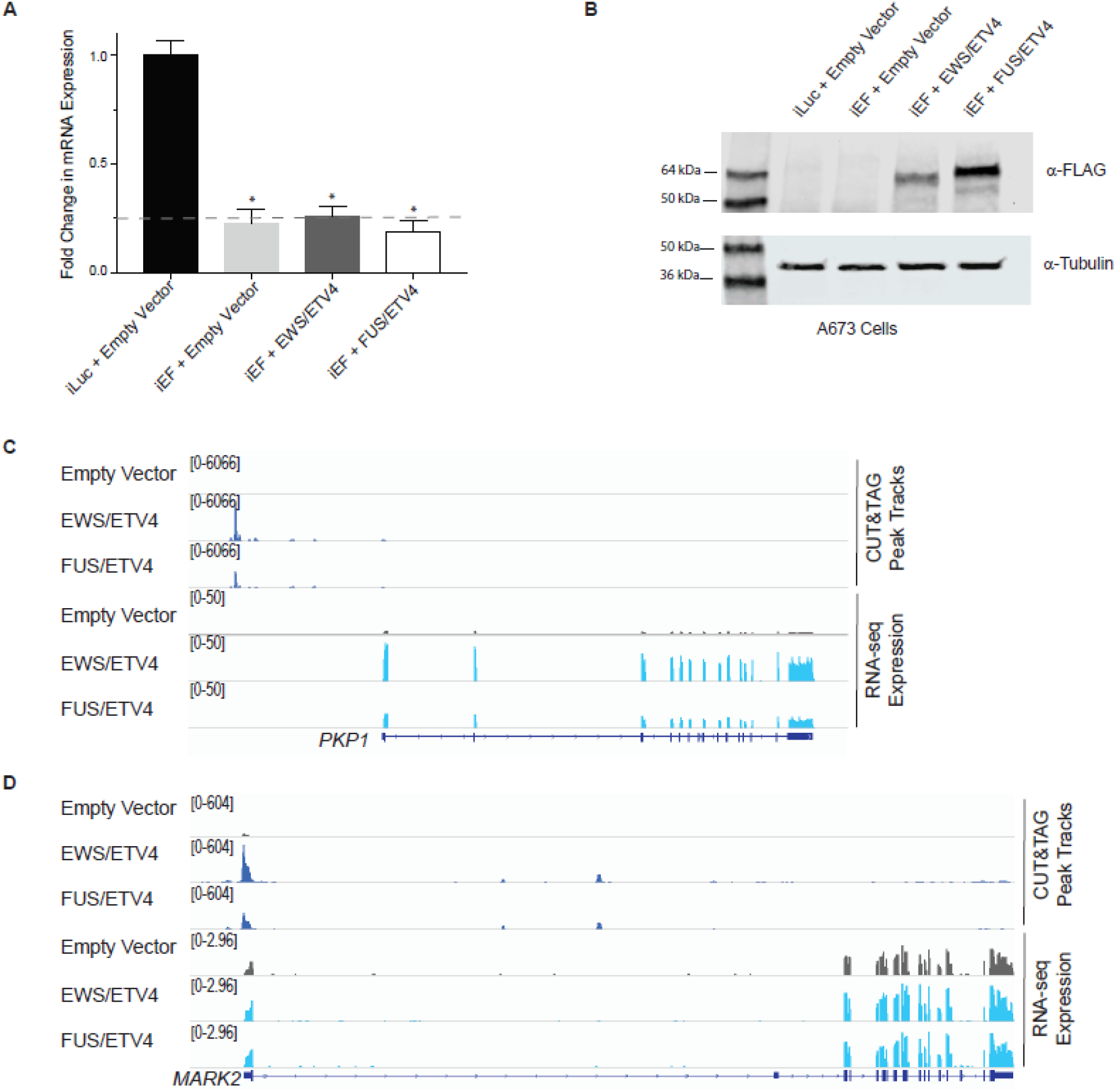
EWS/ETV4 and FUS/ETV4 fusion proteins studied in A673 knock-down/rescue model system. (A) Representative qRT-PCR results of endogenous EWS/FLI mRNA levels in A673 cells harboring the indicated constructs (iLuc: control shRNA; iEF: shRNA targets the 3’UTR of endogenous EWS/FLI). EWS/FLI mRNA values were normalized to RPL30 mRNA control values. Asterisks indicate samples are statistically different as compared to control iLuc + Empty Vector cells (p-value < 0.05, N=1). (B) Western blot of 3xFLAG-tagged EWS/ETV4 and FUS/ETV4 protein expression in A673 cells. Membranes were probed with α-FLAG or α-tubulin (loading control) antibodies. (C-D) CUT&TAG and RNA-sequencing peak tracks visualized for Empty Vector cells (iEF + Empty Vector), EWS/ETV4-containing cells, and FUS/ETV4-containing cells (N=2 for each sample). Example genes include those associated with both microsatellite (*PKP1*) and high-affinity (HA) site (*MARK2*)-regulated genes. Peak track scales are depicted on the left.

**Supplemental Figure 2.**
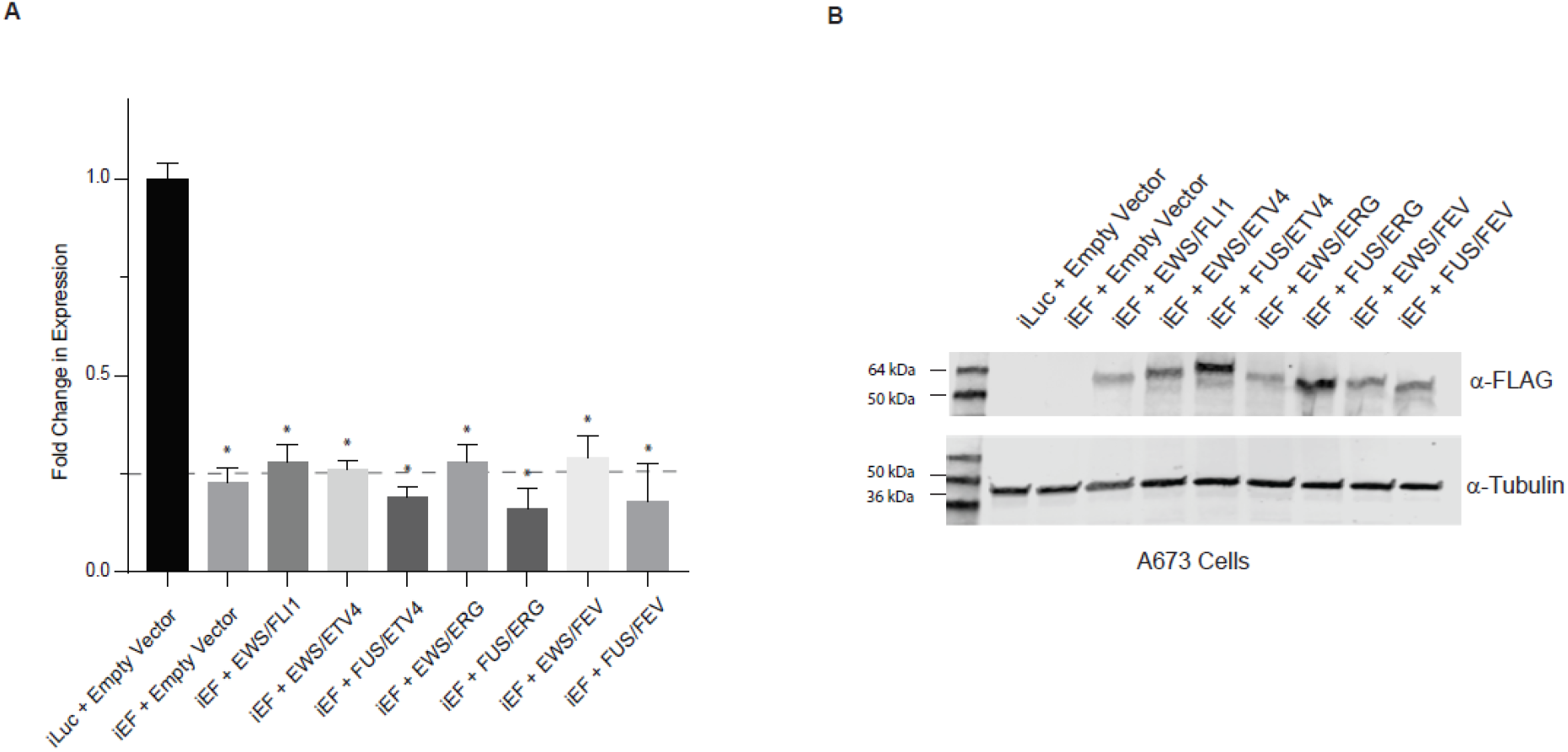
Successful expression of FET/ETS fusion proteins in A673 knock-down/rescue model system. (A) Representative qRT-PCR results determining endogenous EWS/FLI mRNA knock-down in A673 cells. iLuc + Empty Vector cells contain endogenous EWS/FLI mRNA, whereas iEF + Construct samples contain shRNA targeting the 3’UTR of endogenous EWS/FLI mRNA. All samples were normalized to RPL30 mRNA control samples. Statistical significance as compared to iLuc + Empty Vector is indicated by asterisks (p-value < 0.05, N = 1). (B) Western blot analysis demonstrating protein expression of FET/ETS fusion proteins in A673 knock-down/rescue cells. Membranes were probed for protein expression (α-FLAG) and a loading control (α-tubulin).

**Supplemental Figure 3.**
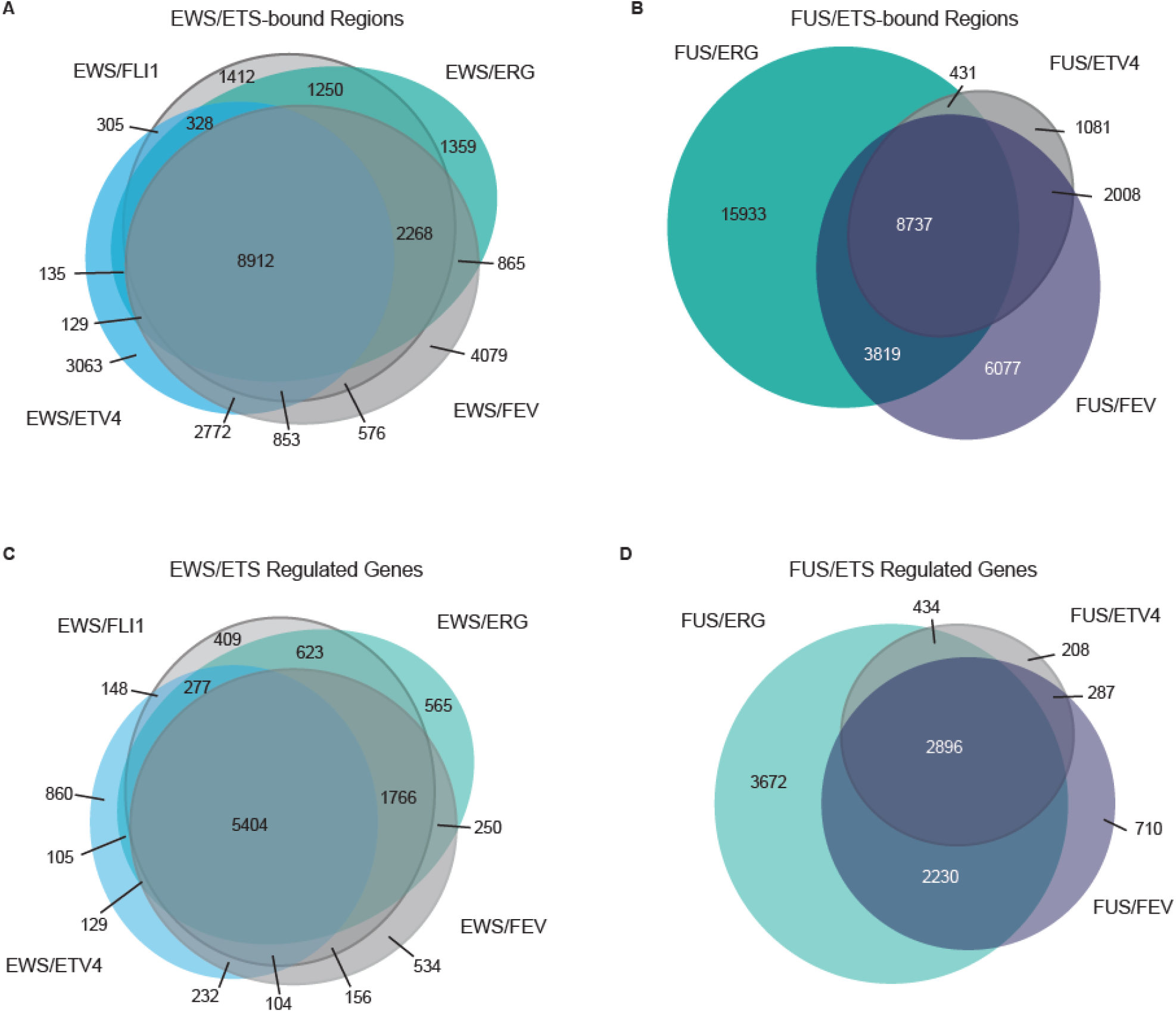
Overlap of EWS/ETS and FUS/ETS fusions reveals similar DNA-binding and transcriptional profiles. (A-B) DNA-bound regions called as significant over background for the (A) EWS/ETS fusions and (B) FUS/ETS fusions were overlapped (N=2 for each sample). Number of individually bound and shared bound regions are indicated in each circle. Significance of overlap: p < 2.2 x 10^−16^. (C-D) Venn diagram analysis depicts significantly regulated genes for (C) EWS/ETS and (D) FUS/ETS fusion proteins. The number of genes regulated by each protein is indicated in the figure (N=2 for each sample). Significance of overlap: p < 2.2 x 10^−16^.

**Supplemental Figure 4.**
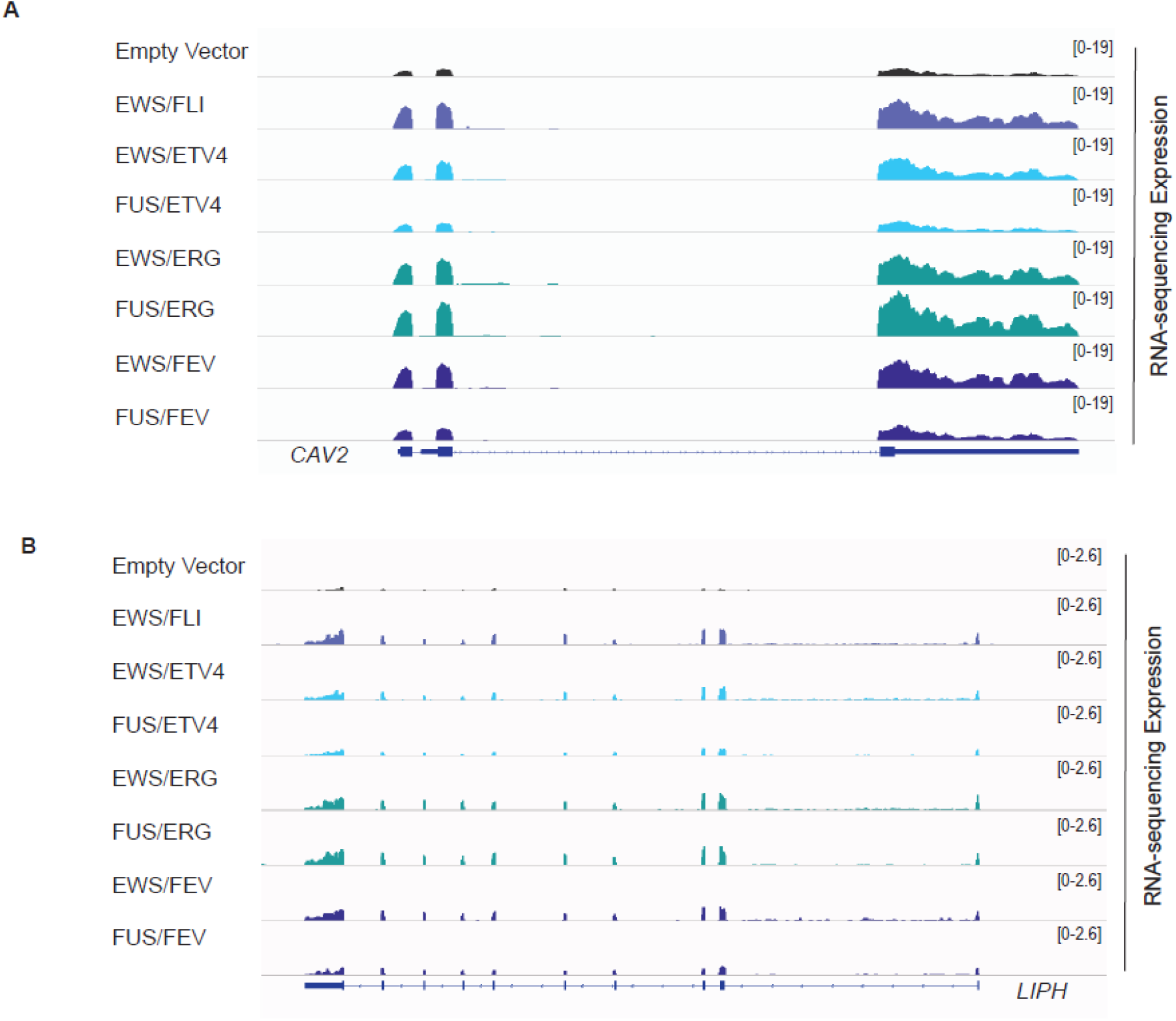
FET/ETS fusion proteins bind at known EWS/FLI response elements in Ewing sarcoma cells. (A-B) Representative peak tracks from CUT&Tag DNA-binding analysis are shown for Empty Vector (iEF + Empty Vector) A673 control cells, as well as A673 knock-down/rescue cells containing each of the FET/ETS fusions listed (N=2 for each sample). Examples of (A) GGAA-microsatellite (*VRK1*) and (B) high-affinity site (*BIRC2*) bound peaks typically associated with EWS/FLI function in Ewing sarcoma cells depicted here. Peak track scales displayed on the right.

**Supplemental Figure 5.**
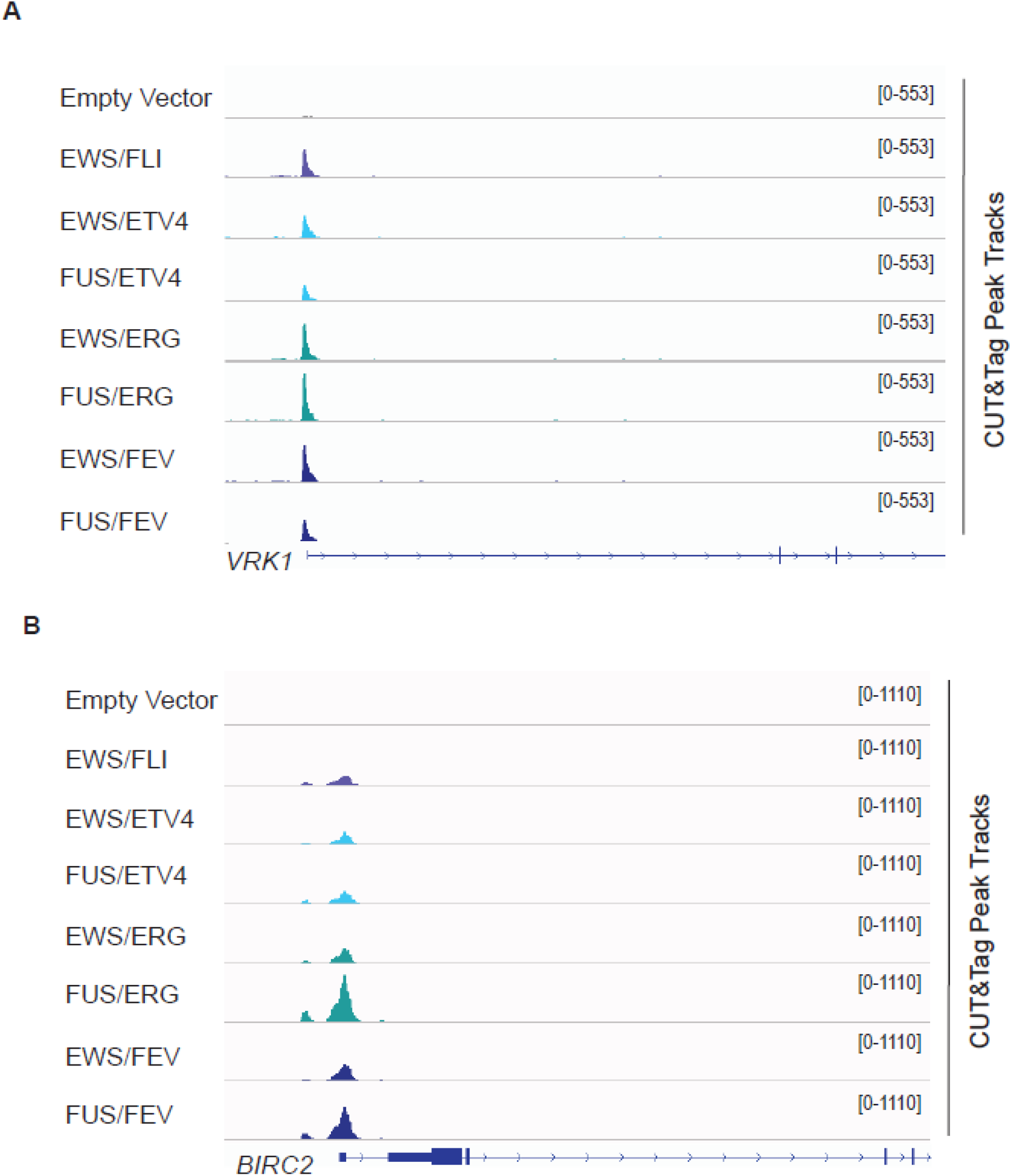
RNA-sequencing analysis reveals FET/ETS fusions regulate genes typically associated with Ewing sarcoma cells. (A-B) Representative tracks of RNA-sequencing expression data from IGV are shown for Empty Vector (iEF + Empty Vector) and rescue samples (iEF + FET/ETS translocation) (N=2 for each sample). Examples of expression data are associated with EWS/FLI regulation in Ewing sarcoma cells via (A) GGAA-microsatellite (*CAV2*) and (B) high-affinity site (*LIPH*). Peak tracks scales are depicted on the right.

**Supplemental Table 1.** Sequence of FET/ETS cDNA Constructs. Sequence of FET/ETS protein-encoding cDNA constructs used for manuscript, including corresponding exon and amino acid information. All translocations studied here directly correlate to translocations identified and reported in the literature found in Ewing sarcoma patient tumors.

| **Translocation** | **FET Exons** | **FET Amino Acids** | **ETS Exons** | **ETS Amino Acids** |
| --- | --- | --- | --- | --- |
| EWS/FLI1<br>EWSR1:NP_001156757.1<br>FLI1: NP_002008.2 | 1-7 | 1-265 | 7-9 | 242-452 |
| EWS/ETV4<br>EWSR1:NP_001156757.1<br>ETV4: NP_001073143 | 1-7 | 1-265 | 9-13 | 271-484 |
| FUS/ETV4<br>FUS: NP_004951.1<br>ETV4: NP_001073143 | 1-10 | 1-355 | 9-13 | 271-484 |
| EWS/ERG<br>EWSR1:NP_001156757.1<br>ERG: NP_891548.1 | 1-7 | 1-265 | 9-12 | 250-479 |
| FUS/ERG<br>FUS: NP_004951.1<br>ERG: NP_891548.1 | 1-7 | 1-255 | 9-12 | 250-479 |
| EWS/FEV<br>EWSR1:NP_001156757.1<br>FEV: NP_059991.1 | 1-10 | 1-347 | 2-3 | 18-238 |
| FUS/FEV<br>FUS: NP_004951.1<br>FEV: NP_059991.1 | 1-10 | 1-355 | 2-3 | 18-238 |

**Supplemental Table 2.**
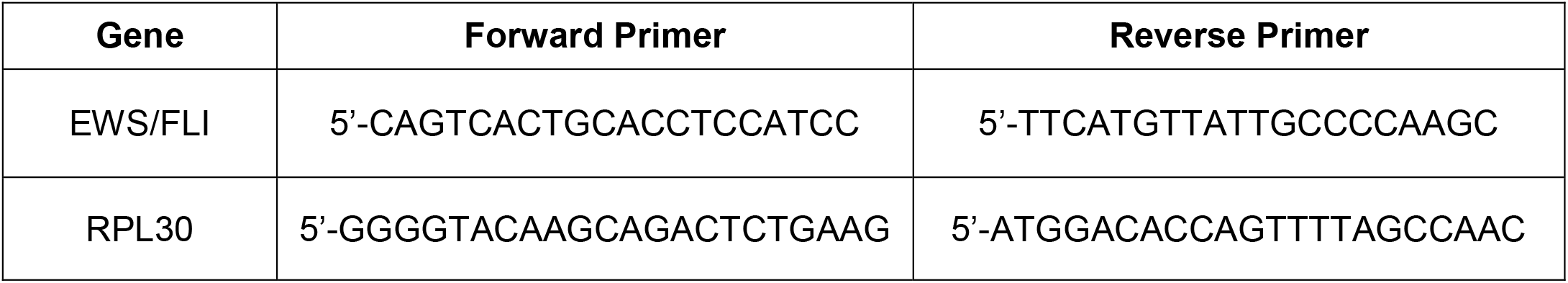
Sequences for primers used in qRT-PCR experiments. Sequences of primers used for qRT-PCR experiments to determine knock-down of endogenous EWS/FLI mRNA. RPL30 is used as a control to normalize data for all samples.

